# Clinical safety and pharmacokinetics of a novel oral niclosamide formulation compared with marketed niclosamide chewing tablets in healthy volunteers: a three-part randomized, double-blind, placebo-controlled trial

**DOI:** 10.1101/2024.05.06.24306928

**Authors:** Niklas Walther, Robert Schultz-Heienbrok, Heino Staß, Victor M. Corman, Nils C. Gassen, Marcel A. Müller, Christian Drosten, Martin Witzenrath, Hweeling Lee, Maximilian G. Posch

## Abstract

**Aim:** Niclosamide is an established anthelmintic substance and a promising candidate for treating cancer, viral infections, and other diseases. However, its solubility in aqueous media is low, and the systemic bioavailability of the commercially available chewing tablet is poor, limiting the use of niclosamide for systemic treatment. A liquid oral formulation using polyethylene glycol 400 was developed and investigated in healthy volunteers to assess safety, tolerability, and pharmacokinetics in comparison to the marketed tablet. (ClinicalTrials.gov: NCT04644705)

**Methods:** The study consisted of three parts: Part A was a double-blind placebo-controlled single ascending dose trial in three dose groups (200, 600, and 1600 mg) with four participants receiving either the investigational niclosamide formulation or placebo (3:1) under fasted and/or fed conditions. Part B was a crossover study comparing 1600 mg investigational niclosamide solution with the marketed 2000 mg chewing tablet in four healthy volunteers. Part C was a double-blind placebo-controlled multiple-dose trial comparing 1200 mg and 1600 mg (verum: placebo 4:2) in two dose groups with six subjects each, who received daily doses for seven days.

**Results:** No serious or severe adverse events occurred. The most frequent adverse events were mild to moderate gastrointestinal reactions. There was also no apparent dependence between drug exposure levels (AUC, Cmax) and the severity and incidence of adverse events detectable. A relevant food effect was observed with a mean AUC_last_ about 2-fold higher in fed condition compared to fasted condition. In Part B, dose-normalized C_max_ and AUC_last_ were similar for niclosamide solution and tablet. Absorption of niclosamide solution was highly variable. Some individuals showed high absorption (C_max_ >2µg/ml) whereas others did absorb only marginally. Importantly, there was no dose linearity in the range of 200 mg – 1600 mg. No signs of relevant systemic drug accumulation after multiple administrations were observed.

**Conclusion:** Overall safety and tolerability observed in healthy subjects were benign. This is also true for individual “high absorbers” (C_max_ >2µg/ml), encouraging further research into niclosamide as a potential therapeutic agent. Galenic optimization, however, will remain challenging as evident from the observed exposure variability and non-linear PK. Non-linearity, if confirmed by additional data, might make niclosamide more suitable for multi-dose rather than high single dose regimens. The observed food effect should also be considered when further investigating systemic niclosamide exposures.

## Introduction

Niclosamide is a chlorinated salicylanilide approved to treat tapeworm infections and marketed for human use since 1962 [1]. In recent years, various studies indicated that niclosamide could also be a therapeutic option for other non-infectious diseases, including malignancies, neurological diseases, metabolic diseases, and skin diseases [2, 3, 4, 5, 6, 7]. Interestingly, recent experimental data also indicate that niclosamide may be effective against SARS-CoV-2 infection through inhibition of BECN1-targeting E3 ligase SKP2 and activation of cellular autophagy processes [8]. A major hurdle in using the potentially beneficial pharmacological properties of the substance is its low aqueous solubility which limits bioavailability and systemic exposure. For COVID-19 as well as for other diseases, relevant plasma levels cannot be reached [9].

Currently, niclosamide (Yomesan©) is marketed as chewing tablets, a formulation developed to hold a topical effect in the gastrointestinal tract. As a result, the bioavailability of Yomesan© is intentionally low. Plasma level concentrations vary between 0.25− 6.0 μg/mL according to the summary of product characteristics (SmPC) for a once daily administration of 2 grams Yomesan©. This variability further limits the value of Yomesan© for clinical use in other indications that require systemic exposure. Other formulations like niclosamide-loaded nanoparticles or inhalable/intranasal formulations are being developed and tested [10, 11, 12, 13, 14].

We developed the drug as a drinking solution consisting of polyethyleneglycol 400 g/mol (PEG 400) as a solvent for niclosamide, aiming to increase intestinal absorption and to ensure more reproducible uptake [15, 16]. We hypothesized that the niclosamide solution would reduce the high variability of plasma levels and increase systemic availability, making it more suitable for the clinical treatment of systemic diseases.

The primary objectives of this study were to evaluate the safety, tolerability, and pharmacokinetic (PK) profiles of single ascending doses (SAD) of the niclosamide solution in healthy volunteers. The secondary objectives were to assess the safety, tolerability, and PK profiles of multiple ascending doses (MAD) and to evaluate the PK of a single dose of the niclosamide solution under fed and fasted conditions. Furthermore, the relative bioavailability of the niclosamide solution compared to the marketed chewing tablet was assessed. We utilized an adaptive trial design that enabled us to adjust the dose based on the safety and PK data obtained throughout the study (S1 Protocol).

## Methods

### Study Design

This trial was a single-centre, 3-part (Parts A, B, C) randomized phase 1 trial to investigate the safety and pharmacokinetics of an investigational niclosamide solution in 28 healthy volunteers. A Data Safety Monitoring Board (DSMB) continuously monitored subject safety and decided on dose escalation in part A and initiation of parts B and C.

Part A was a randomized, double-blind, placebo-controlled SAD study with three consecutive dose cohorts including 4 participants (verum: placebo 3:1). Furthermore, subjects enrolled in cohort A3 received the drug first under fasted and later under fed conditions after a washout period to investigate differences in bioavailability with regard to the fasted status.

Part B consisted of one randomized, open-label, two-sequence, two-period crossover cohort comparing the treatment achieving the highest exposure tested to be safe and tolerable of the niclosamide solution in part A with the marketed chewing tablets.

Part C was a randomized, double-blinded, placebo-controlled multiple ascending dose study investigating the safety and pharmacokinetics of the niclosamide solution over a treatment period of seven days. This part consisted of two dose groups.

### Ethical conduct

The study was conducted at the Phase 1 Study Unit of Charite Research Organisation GmbH in Berlin, Germany following Good Clinical Practices (ICH-GCP) and the Declaration of Helsinki. An independent Ethics Committee (Landesamt für Gesundheit und Soziales [LAGeSo]) and the German competent authority (Bundesinstitut für Arzneimittel und Medizinprodukte [BfArM]) reviewed and approved the study before initiation. The study is registered with ClinicalTrials.gov, NCT04644705, and EU Clinical Trials Register, EudraCT 2020-003451-15.

### Participants

The study was conducted from November 17, 2020, to May 3, 2021. Healthy male and female volunteers confirmed by medical history, physical examination, vital signs, and safety laboratory at screening between 18 and 45 years understanding the content of informed consent were eligible to participate in the trial. Key exclusion criteria were clinically relevant medical conditions and pregnant or lactating women. Before any screening activities, volunteers gave written informed consent to participate in the study (see S1 Protocol for the complete list of inclusion and exclusion criteria).

### Randomization and blinding

Niclosamide investigational solution, chewing tablets, and placebo solution were provided by Bayer AG, Germany, prepared and dispensed by an unblinded pharmacist of the clinical trial supply (CTS) team at Charité Research Organization GmbH, and subsequently administered by a blinded investigator. The placebo for the solution contained diluent (PEG) and levomenthol as flavour. Levomenthol was also part of the niclosamide solution, thus providing the same taste and volume as the placebo solution. Since the investigational solution and placebo were different in colour, CTS prepared the medicinal product in identical syringes covered with a non-transparent foil to sustain blinding. The administration of investigational medicinal product (IMP, which could be either treatment or placebo) was randomized in a 3 (treatment): 1 (placebo) in part A (SAD), 4 (treatment):0 in part B (cross-over treatment) and 4 (treatment) 2 (placebo) in Part C. Eligible participants were randomized according to a randomization list generated with SAS® 9.4 (SAS Institute Inc., USA). The randomization list was not accessible by blinded staff; however, sealed emergency code break envelopes were available for the investigators in case of emergency. The IMP was provided to the investigator by the pharmacist in a blinded manner. A sentinel dosing approach was used in part A, meaning that the first subject in each cohort received an open-label administration of the niclosamide solution, followed by three double-blinded subjects in each cohort. In parts A and C, investigators, study staff, and participants were blinded throughout the trial.

### Study Interventions

In part A, the IMP was administered on study day 1 under fasted conditions in cohorts A1 (200 mg niclosamide or placebo) and A2 (600 mg niclosamide or placebo) after safety and tolerability of 200 mg have been confirmed by the DSMB. Participants were fasting for at least 9 hours before and at least 4 hours after IMP intake. In cohort A3 (1600 mg or placebo), the IMP was first administered once in fasted condition and then, after a washout phase of 16 to 18 days, administered again in the same subjects under fed condition, at the same dose level. Under fed condition, a high-fat, high-calorie breakfast (Guidance for Industry: Food-Effect Bioavailability and Fed Bioequivalence Studies, 2001) following an overnight fast of at least 9 hours starting 30 minutes before treatment was consumed. Water intake was not permitted from 2 hours before to 2 hours after receiving the IMP. In all parts, administration was once daily (QD), orally in the morning in an upright position with 240 ml of non-sparkling water. All subjects were informed about dietary/activity requirements and restrictions and adhered to them.

Parts B and C were conducted under fed conditions, as cohort A3 showed that systemic exposure of the niclosamide solution was higher and safe with prior food intake. In part B, the highest tolerated and safe dose of niclosamide investigational solution in part A was compared with the approved dose of 2000 mg of the marketed niclosamide chewing tablet. Each subject received treatment with 1600 mg niclosamide as an oral solution and 2000 mg niclosamide as a chewing tablet on study days 1 and 3 (the first day of period 2). On day 1 of treatment period 1, either a single dose of the solution or the chewing tablet was administered. After a one-day wash-out between each period, the other treatment variant was administered on day 3.

Part C consisted of two dose groups with six subjects each. Each subject in cohort 1 received treatment with 1200 mg niclosamide or placebo and subjects enrolled in cohort 2 received 1600 mg niclosamide or placebo once daily orally from study day 1 to 7 under fed condition.

### Bioanalytical methods

In part A and part B, blood samples for measuring plasma niclosamide concentrations were collected predose and at 0.5, 1.0, 1.5, 2, 4, 6, 8, 12 hours, post-dose on day 1, as well as on day 2, 24 hours after dose administration. In part B, plasma levels were also measured on day 3 at the same timepoints. In part C, PK samples were collected at predose, and 0.5, 1.0, 1.5, 2, 4, 6, 8, 12 hours, and 24 h post-dose after first and last dose administration (study days 1 and 7), as well as predose on days 2 to 6 (trough levels). Blood was collected and centrifuged; plasma was separated for bioanalytical analyses, frozen within 85 minutes of collection, and stored at -80°C until shipment. BIOTEZ Berlin-Buch GmbH measured niclosamide concentration by HPLC method.

#### Pharmacokinetic evaluation

Pharmacokinetics (PK) were evaluated using Phoenix WinNolin® software version 8.3 (Certara, USA). The individual and mean plasma level versus time curves were evaluated using non-compartmental method. The AUC_last_ was calculated as the area under the plasma concentration-time curve from time zero up to the time of the last quantifiable concentration calculated by linear up / log down method. Food effect was evaluated using non-linear mixed effect modelling.

### Safety assessments

Safety was assessed by clinical laboratory tests, physical examinations, 12-lead electrocardiograms (ECG), vital signs, and monitoring of adverse events (AEs). At screening and each visit to the research unit, safety assessments were performed. Investigators assessed subjects throughout the study for the occurrence of AEs and their severity and their relationship to the IMP. The DSMB continuously monitored subjects’ safety and made decisions on dose escalation. Stopping rules included, occurrence of a related SAE in at least one subject, related severe AEs in two subjects in the same cohort or relevant and confirmed safety laboratory deviations in two or more subjects. Dose escalation would have been stopped if plasma levels of the niclosamide solution exceed specific thresholds (approximately 1.6 µg/ml) at 8 hours post-dosing in all subjects. For the high dose group of 1600 mg in Part C, dosing may have been discontinued if moderate to severe gastrointestinal side effects occurred.

### Outcomes

Primary safety outcome was monitored by the number of treatment-emergent (serious) Adverse Events (TEAE), and the recording of vital signs, ECG, and safety laboratory parameters. Secondary outcomes included the effect of food on rate and extent of absorption and various PK parameters after multiple dosing. PK profiles included the maximum plasma concentration of niclosamide (C_max_) and the area under the plasma concentration time curve from predose until the last detectable concentration of niclosamide (AUC_last_). These parameters were also used to assess relative bioavailability of the solution as compared to the tablets.

### Sample size

The number of subjects was considered adequate to investigate safety and was not based on considerations of statistical power due to the exploratory and descriptive nature of the study. In parts A and B, each dose group consisted of four subjects, with three subjects randomly assigned to receive niclosamide solution. In part C, each dose group included six subjects, with four subjects randomized to receive the solution.

### Statistical methods

The primary objective was to evaluate the safety and PK of single ascending doses of the oral solution. The data obtained were descriptive since the trial was not designed for inferential statistics. A safety analysis was done in all participants who received at least one dose of study treatment. AEs and SAEs were coded according to the Medical Dictionary for Regulatory Activities (MedDRA) and recorded by system organ class (SOC) and preferred term. ECGs, Physical examinations, and clinical laboratory abnormalities were summarized.

Descriptive statistics were calculated for the plasma concentration of niclosamide at each applicable time point specified and for the derived plasma PK parameters (C_max_, AUC_last_, C8h, C12h, C24h, t_max_, t_1/2_). Geometric means (GM) and standard deviations (SD) were described for C_max_, AUC and t_1/2_ . Medians and ranges were used for T_max_.

The food effect evaluation in part A and the relative bioavailability compared to the chewing tablets in part B were performed using a linear fixed-effect model containing fixed effects for sequence, treatment, period, and subjects within sequence for log-transformed C_max_, and AUC_last_, respectively. Only subjects with evaluable data for both periods were included.

All outcomes were analyzed based on per-protocol analysis (all subjects who completed the trial without major protocol deviations).

## Results

### Subjects

Twenty-eight healthy subjects were enrolled in part A (12 subjects), part B (4 subjects), and part C (12 subjects), all completed the study according to protocol. Of all participants, 21 received niclosamide and 7 received placebo (Fig 1). The demographic and clinical characteristics of the subjects are summarised in Table 1.

**Fig 1.**
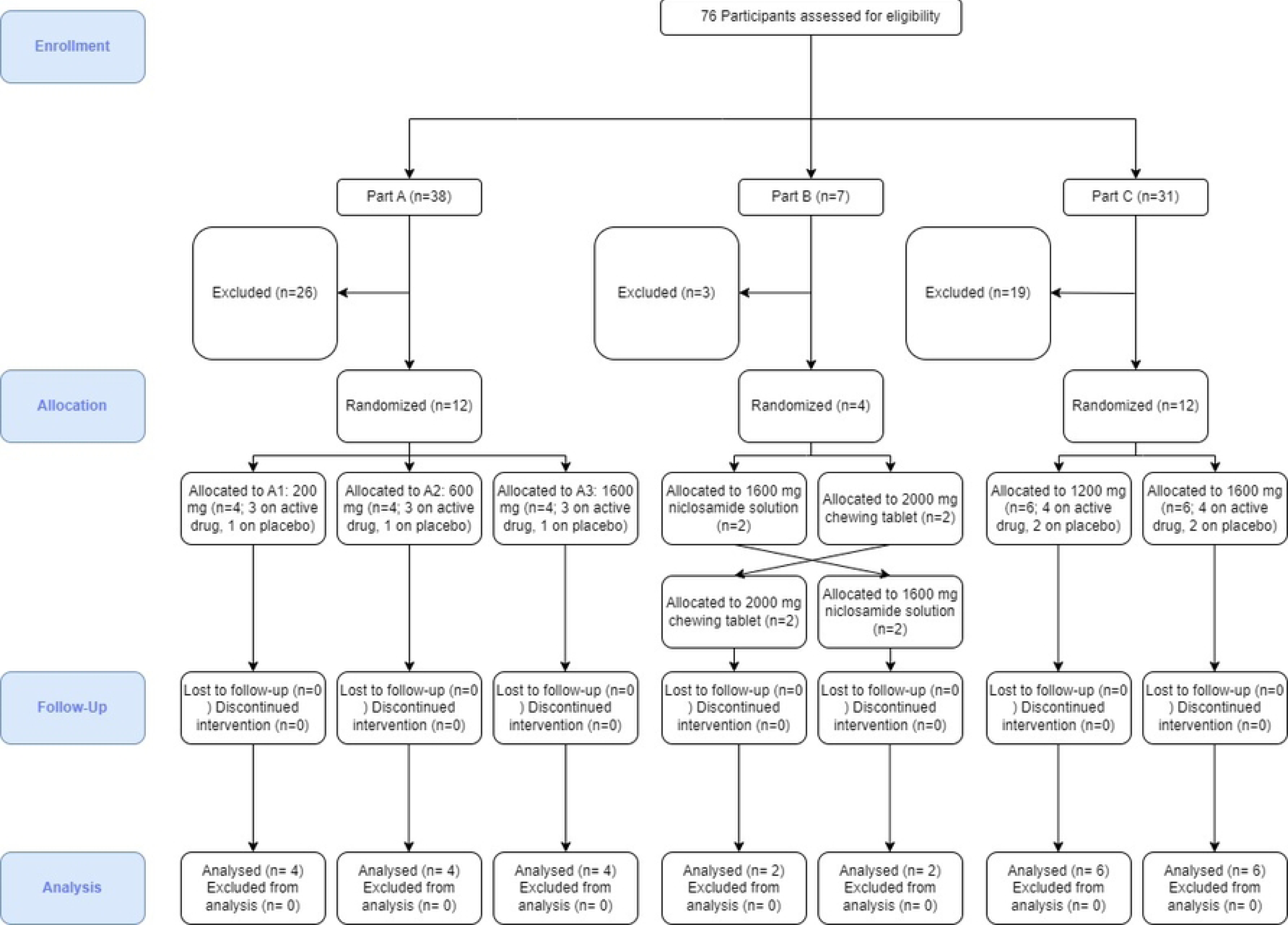
Study CONSORT flowchart.

**Table 1:** Demographic and Baseline Characteristics of Participants.

| Characteristic | Part A (200 mg) | Part A (600 mg) | Part A (1600 mg) | Part A (Placebo) | Part B | Part C (1200 mg) | Part C (1600 mg) | Part C (Placebo) |
| --- | --- | --- | --- | --- | --- | --- | --- | --- |
| Gender: Female (n, %) | 3 (100.0%) | 3 (100.0%) | 3 (100.0%) | 3 (100.0 %) | 4 (100.0 %) | 4 (100.0 %) | 4 (100.0 %) | 4 (100.0 %) |
| Age: Mean, SD [y] | 28.7, 9.7 | 28.0, 2.0 | 23.7, 2.5 | 31.0, 7.8 | 28.8, 2.9 | 30.3, 5.7 | 33.8, 6.4 | 32.8, 6.3 |
| Race: White/Caucasian, (n, %) | 3 (100.0%) | 3 (100.0%) | 3 (100.0%) | 3 (100.0 %) | 4 (100.0 %) | 4 (100.0 %) | 4 (100.0 %) | 4 (100.0 %) |
| Smoking History: Current (n, %) | 0 (0.0%) | 0 (0.0%) | 0 (0.0%) | 0 (0.0%) | 3 (75.0%) | 1 (25.0%) | 0 (0.0%) | 1 (25.0%) |
| Smoking History: Former (n, %) | 2 (66.7%) | 0 (0.0%) | 0 (0.0%) | 1 (33.3%) | N/A | 1 (25.0%) | 3 (75.0%) | 1 (25.0%) |
| Smoking History: Never (n, %) | 1 (33.3%) | 3 (100.0%) | 3 (100.0%) | 2 (66.7%) | 1 (25.0%) | 2 (50.0%) | 1 (25.0%) | 2 (50.0%) |
| Weight: Mean, SD [kg] | 58.07, 0.8 | 65.77, 13.23 | 61.0, 2.15 | 64.63, 14.51 | 70.50, 16.02 | 63.70, 16.56 | 70.75, 7.10 | 62.45, 7.31 |
| Height: Mean, SD [cm] | 168.3, 3.1 | 173.7, 3.8 | 170.7, 2.1 | 165.7, 11.6 | 169.5, 6.7 | 166.8, 5.7 | 164.8, 4.4 | 164.8, 5.1 |
| BMI: Mean, SD [kg/m2] | 20.47, 0.47 | 21.7, 3.36 | 20.93, 1.01 | 23.33, 1.97 | 24.33, 3.59 | 22.70, 4.65 | 26.00, 1.26 | 23.08, 3.06 |

SD = standard deviation; n= number of subjects

### Safety outcome

No serious or severe AEs occurred throughout the study, and no clinically relevant changes from baseline were observed in any subject in physical examination, vital signs, ECGs, or safety laboratory parameters. None of the stopping criteria were met in any subject during the trial.

During the trial, 58 TEAEs occurred in 21 subjects, meaning 75% of all subjects experienced at least one or more AEs. Of these, 54 were of mild intensity and four of moderate intensity. Under “gastrointestinal disorders”, 45 (78 %) TEAEs were grouped, with “diarrhoea” (including fluid stool), (16), “nausea” (12), and “vomiting” (4) as the most frequent events. For this System Organ Class (SOC), events were experienced by 14 out of 21 subjects (67%) who received treatment and by 6 out of 10 (60%) subjects who received placebo. Overall, seven (58%) subjects in part A, two (50%) subjects in part B and twelve (100%) subjects in part C reported at least one AE. The occurrence of suspected drug-related AEs was 18 in 7 subjects (58.2%) in part A, 3 AEs in 2 (50%) subjects in part B, and 28 AEs in 12 (100%) subjects in part C.

In part A, only one subject 1/9 (11%) in the treatment cohort and none in the placebo cohort experienced a moderate TEAE “abdominal pain”, which was considered to be related to the IMP by the investigator. All other AEs were of mild intensity (Table 2).

**Table 2.**
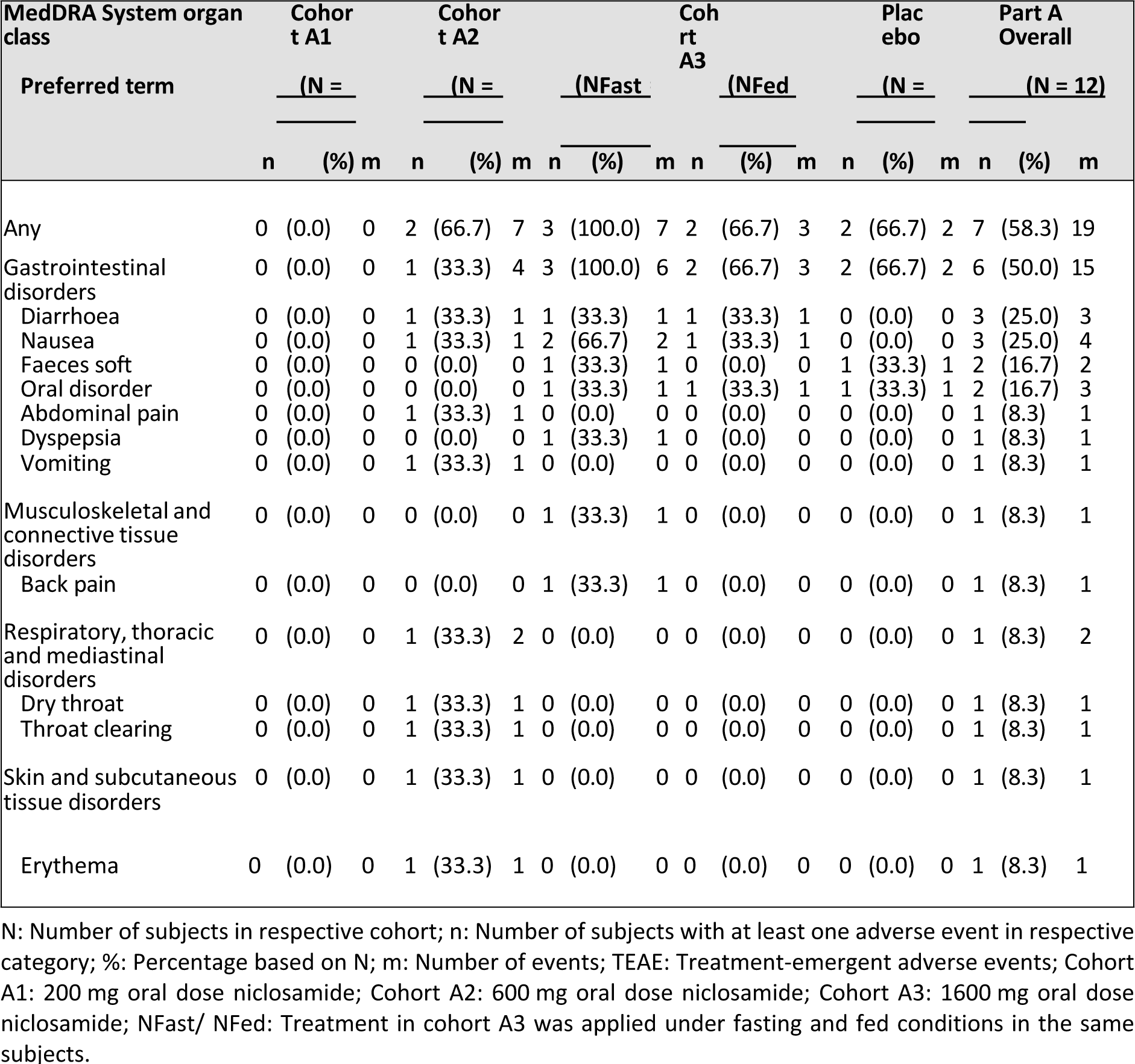
Adverse Events by MedDRA SOC and PT – Part A.

In part B, the three AEs, “nausea,” “diarrhoea” and “vomiting,” occurred on the same day after ingestion of the niclosamide solution and were judged as related, whereas no AEs were observed after intake of the chewing tablet (Table 5).

**Table 5.** Adverse Events by MedDRA SOC and PT – Part B.

| MedDRA System organ class<br>Preferred term | Solution 1600mg<br>(N = 4) |  |  | Chewing tablet<br>2000mg<br>(N = 4) |  |  | Part B Overall<br>(N = 4) |  |  |
| --- | --- | --- | --- | --- | --- | --- | --- | --- | --- |
|  | n | (%) | m | n | (%) | m | n | (%) | m |
| Any | 2 | (50.0) | 3 | 0 | (0.0) | 0 | 2 | (50.0) | 3 |
| Gastrointestinal disorders | 2 | (50.0) | 3 | 0 | (0.0) | 0 | 2 | (50.0) | 3 |
| Diarrhoea | 1 | (25.0) | 1 | 0 | (0.0) | 0 | 1 | (25.0) | 1 |
| Nausea | 1 | (25.0) | 1 | 0 | (0.0) | 0 | 1 | (25.0) | 1 |
| Vomiting | 1 | (25.0) | 1 | 0 | (0.0) | 0 | 1 | (25.0) | 1 |

N: Number of subjects in respective cohort; n: Number of subjects with at least one adverse event in respective category; %: Percentage based on N; m: Number of events; TEAE: Treatment-emergent adverse events; Part B was a crossover design.

In part C, all subjects of the 1200 mg group, the 1600 mg group, and the placebo group experienced TEAEs. Most TEAEs were mild in severity, except for three moderate TEAEs, specifically two incidences of “diarrhoea” experienced in two participants in each of the 1200 mg group and placebo group, and “nausea” reported in one participant from the 1600 mg group. Three events of nausea and one event of vomiting were rated as not related, as the subjects could credibly attribute them to the high-fat breakfast. A dose-dependent increase in TEAEs among subjects in part C was not observed (Table 6). All subjects had at least one or more events.

**Table 6.** Adverse Events by MedDRA SOC and PT – Part C.

|  | n | (%) | m | n | (%) | m | n | (%) | m | n | (%) | m |
| --- | --- | --- | --- | --- | --- | --- | --- | --- | --- | --- | --- | --- |
| Any | 4 | (100.0) | 13 | 4 | (100.0) | 14 | 4 | (100.0) | 9 | 12 | (100.0) | 36 |
| Gastrointestinal disorders | 4 | (100.0) | 9 | 4 | (100.0) | 11 | 4 | (100.0) | 7 | 12 | (100.0) | 27 |
| Diarrhoea | 4 | (100.0) | 4 | 4 | (100.0) | 5 | 3 | (75.0) | 3 | 11 | (91.7) | 12 |
| Nausea | 2 | (50.0) | 2 | 4 | (100.0) | 4 | 1 | (25.0) | 1 | 7 | (58.3) | 7 |
| Flatulence | 1 | (25.0) | 1 | 0 | (0.0) | 0 | 2 | (50.0) | 2 | 3 | (25.0) | 3 |
| Vomiting | 0 | (0.0) | 0 | 2 | (50.0) | 2 | 0 | (0.0) | 0 | 2 | (16.7) | 2 |
| Dry mouth | 1 | (25.0) | 1 | 0 | (0.0) | 0 | 0 | (0.0) | 0 | 1 | (8.3) | 1 |
| Faeces soft | 0 | (0.0) | 0 | 0 | (0.0) | 0 | 1 | (25.0) | 1 | 1 | (8.3) | 1 |
| Rectal haemorrhage | 1 | (25.0) | 1 | 0 | (0.0) | 0 | 0 | (0.0) | 0 | 1 | (8.3) | 1 |
| Nervous system disorders | 2 | (50.0) | 2 | 1 | (25.0) | 2 | 1 | (25.0) | 1 | 4 | (33.3) | 5 |
| Headache | 2 | (50.0) | 2 | 1 | (25.0) | 2 | 1 | (25.0) | 1 | 4 | (33.3) | 5 |
| General disorders and administration site conditions | 1 | (25.0) | 1 | 0 | (0.0) | 0 | 1 | (25.0) | 1 | 2 | (16.7) | 2 |
| Fatigue | 0 | (0.0) | 0 | 0 | (0.0) | 0 | 1 | (25.0) | 1 | 1 | (8.3) | 1 |
| Puncture site pain | 1 | (25.0) | 1 | 0 | (0.0) | 0 | 0 | (0.0) | 0 | 1 | (8.3) | 1 |
| Infections and infestations | 0 | (0.0) | 0 | 1 | (25.0) | 1 | 0 | (0.0) | 0 | 1 | (8.3) | 1 |
| Rhinitis | 0 | (0.0) | 0 | 1 | (25.0) | 1 | 0 | (0.0) | 0 | 1 | (8.3) | 1 |
| Respiratory, thoracic and mediastinal disorders | 1 | (25.0) | 1 | 0 | (0.0) | 0 | 0 | (0.0) | 0 | 1 | (8.3) | 1 |
| Cough | 1 | (25.0) | 1 | 0 | (0.0) | 0 | 0 | (0.0) | 0 | 1 | (8.3) | 1 |

N: Number of subjects in respective cohort; n: Number of subjects with at least one adverse event in respective category; %: Percentage based on N; m: Number of events; TEAE: Treatment-emergent adverse events;

### Pharmacokinetic Results

#### Plasma concentrations niclosamide solution

C_max_ and AUC_last_ showed a non-linear absorption behaviour of the niclosamide solution, except for AUC_last_ in cohorts A1 to A2 suggesting no deviation from dose proportionality in the range of 200 to 600 mg. The mean concentration-time profiles of each cohort remained similar throughout the study, showing a rapid increase and decline in plasma concentration (Fig 2). Variability continued to be very high across dose groups for C_max_ and AUC_last_, especially in fasting conditions and for higher doses. The highest values for C_max_ and AUC_last_ were 2.77 µg/ml and 19.15 µg*h/ml, respectively, and were achieved with the solution under fed conditions.

**Fig 2.**
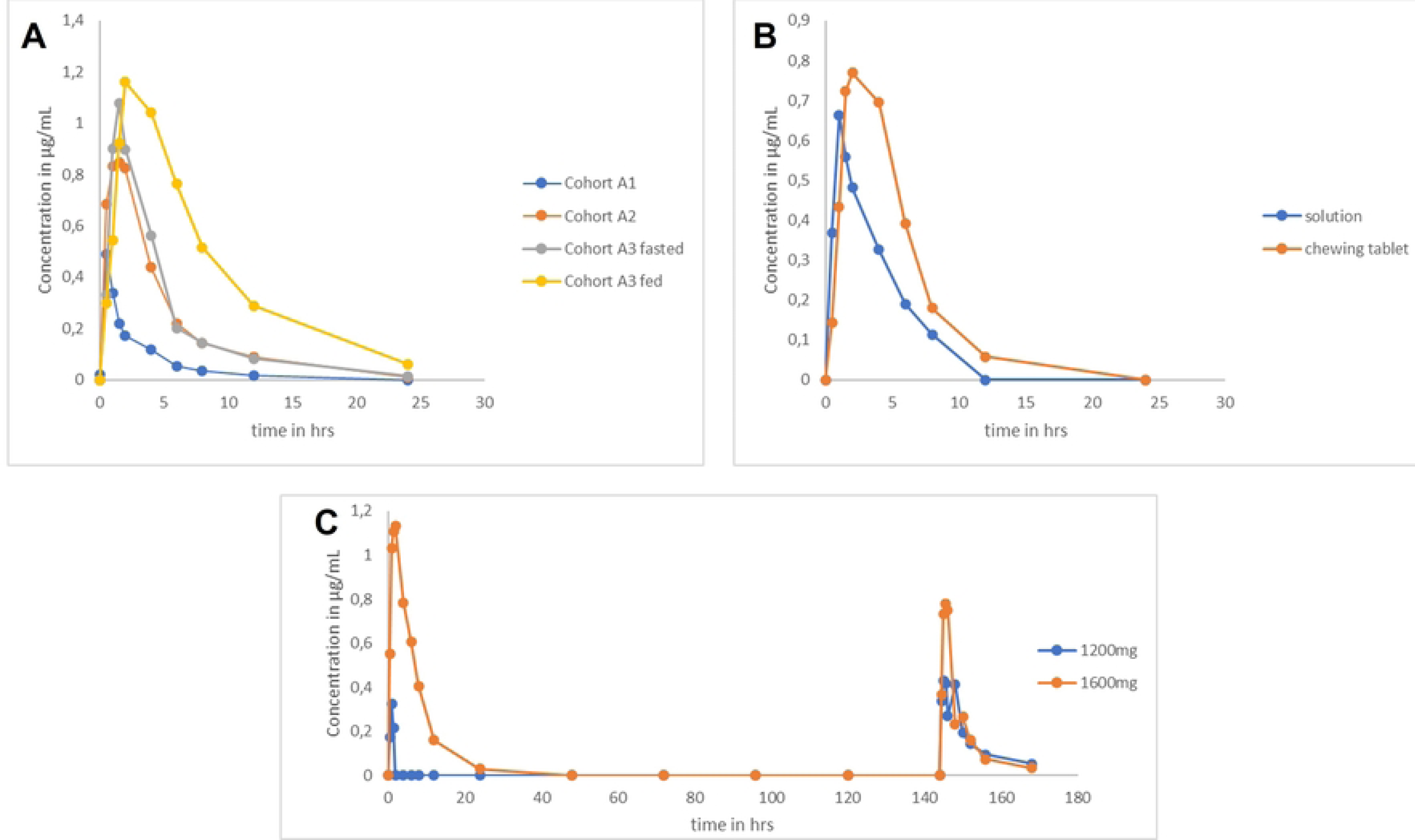
Geometric Mean Niclosamide Plasma-Concentration vs Time Curves following oral administration per cohort. (A) Single ascending doses (200 mg - Cohort A1, 600 mg - A2, 1600 mg - A3) under fasted conditions (A3 also in fed state). (B) 1600 mg solution and 2000 mg tablets under fed conditions. (C) 1200 mg and 1600 mg solution over 7 days in fed state.

In the SAD part A, median T_max_ ranged from 0.5 to 1.5 h across (fasted) cohorts and trended toward longer times with increasing doses. Individual plasma concentrations followed first-order kinetics, declining exponentially over elimination phase. . Half-lives (GM) increased with increasing dose levels, from 3.43 h for the 200 mg dose to 4.15 h for the 1600 mg dose. C_max_ (GM) increased 2.0 -fold from A1 to A2and and only 1.3-fold from A2 to A3. AUC_last_ (GM) levels for the same cohorts increased 3.4 -fold (A1 to A2) and 1.2-fold (A2 to A3) (Table 7).

**Table 7:** Niclosamide Pharmacokinetics for ascending single oral doses in Fasted and Fed conditions - Part A.

| Parameters | Cohort A1 fasted | Cohort A2 fasted | Cohort A3 fasted | Cohort A3 fed |
| --- | --- | --- | --- | --- |
|  | 200 mg | 600 mg | 1600 mg | 1600 mg |
|  | n=3 | n=3 | n=3 | n=3 |
| $C_{max}$, µg/mL | 0.41 [2.31] | 0.84 [1.53] | 1.08 [4.11] | 1.28 [1.64] |
| $AUC_{last}$, µg*h/mL | 1.17 [1.09] | 3.99 [1.71] | 4.86 [3.38] | 9.9 [1.78] |
| $t_{max}$, h | 0.5 [0.5-1.0] | 1.5 [1.5-2.0] | 1.5 [1.5-1.5] | 4.0 [2.0-4.0] |
| C8h, µg/mL | 0.03 [2.40] | 0.12 [1.94] | 0.14 [2.48] | 0.52 [1.88] |
| C12h, µg/mL | BQL | 0.07 [2.31] | 0.08 [3.19] | 0.29 [1.49] |
| C24h, µg/mL | BQL | BQL | 0.01 [1.88] | 0.06 [1.32] |
| $t_{1/2}$, h | 3.43 [2.95] | 4.15 [1.15] | 4.77 [1.14] | 5.28 [1.34] |

$C_{max}$ : Max plasma concentration; $AUC_{last}$ : Area under plasma conc-time curve to last quantifiable conc; $t_{max}$ : Time to reach $C_{max}$ (median [range]); C8h, C12h, C24h: Plasma conc at 8, 12, 24 hrs post-dose; $t_{1/2}$ : Terminal half-life; n: Subjects; BQL: Below quantifiable limit. Geometric mean and standard deviation presented.

#### Food effect

A relevant effect of food intake on the absorption of niclosamide was observed as the median t_max_ was prolonged under fed conditions compared to fasted conditions (4 vs. 1.5 h). T_1/2_ was only slightly higher fasted compared to fed conditions (4.77 vs. 5.28 h) (Fig 2A).

GM C_max_ was 1.2-fold higher for fed than fasting, and the GM AUC_last_ doubled for fed compared to fasting conditions with lower variability (9.90 [1.78] vs. 4.86 [3.38] µg*h/mL) (Table 7) suggesting a substantially increased extent of absorption when niclosamide is administered after a high-fat, high-calorie meal.

#### Relative Bioavailability of niclosamide solution

In part B, the median t_max_ was 1.5 h for the chewing tablet compared with 1 h for the solution, in line with expectations for each formulation. T_1/2_ was slightly shorter for the tablet cohort than the solution cohort and considerably shorter than in the Fed part A3 (Table 8). Notably, the variability in PK parameters was lower for the chewing tablet.

**Table 8.** Comparison of Niclosamide PK parameters for solution and chewing tablet formulations under Fed conditions - Part B.

| PK-Parameters | Niclosamide solution<br>1600 mg fed<br>n=4 | Niclosamide chewing tablets<br>2000 mg fed<br>n=4 |
| --- | --- | --- |
| $C_{\max}$, $\mu\text{g}/\text{mL}$ | 0.78 [3.00] | 0.91 [1.45] |
| $AUC_{\text{last}}$, $\mu\text{g}\cdot\text{h}/\text{mL}$ | 3.47 [2.99] | 5.16 [1.22] |
| $C_{\max}/D$, $\mu\text{g}/\text{mL}$ | 0.49 [3.00] | 0.45 [1.45] |
| $AUC_{\text{last}}/D$, $\mu\text{g}\cdot\text{h}/\text{mL}$ | 2.17 [2.99] | 2.58 [1.22] |
| $t_{\max}$, h | 1.0 [0.5-2.0] | 1.5 [1.0-4.0] |
| $C_8\text{h}$, $\mu\text{g}/\text{mL}$ | 0.12 [2.29] | 0.18 [1.88] |
| $C_{12\text{h}}$, $\mu\text{g}/\text{mL}$ | BQL | 0.06 [5.44] |
| $C_{24\text{h}}$, $\mu\text{g}/\text{mL}$ | BQL | BQL |
| $t_{1/2}$, h | 3.99 [1.49] | 3.66 [1.93] |

C<sub>max</sub>: Max plasma concentration; AUC<sub>last</sub>: Area under plasma conc-time curve to last quantifiable conc; C<sub>max</sub>/D, AUC<sub>last</sub>/D: C<sub>max</sub>, AUC<sub>last</sub> normalized to dose; t<sub>max</sub>: Time to reach C<sub>max</sub> (median [range]); C<sub>8h</sub>, C<sub>12h</sub>, C<sub>24h</sub>: Plasma conc at 8, 12, 24 hrs post-dose; t<sub>1/2</sub>: Terminal half-life; n: Subjects; D: Dose (1.6g solution, 2g tablet); BQL: Below quantifiable limit. Geometric mean and standard deviation presented.

Dose-normalized AUC_last_ and C_max_ were comparable between niclosamide solution and niclosamide chewing tablets (C_max_/D 0.49 [3.00] vs. 0.45 [1.45] µg/mL per g and AUC_last_/D 2.17 [2.99] vs. 2.58 [1.22] µg*h/mL per g) indicating that the solution did not lead to an increased bioavailability.

#### Plasma concentrations multiple dosing

In the MAD part C, the GM of t_1/2_ increased in dose group 1200 mg from day 1 (5.26 h) to day 7 (5.76 h). On the other hand, the GM t_1/2_ in dose group 1600 mg was overall slightly shorter than in the 1200 mg group, with 4.15 h on day 1 and 5.49 h on day 7 (Table 9).

**Table 9.** Niclosamide PK parameters for multiple doses of 1200 mg and 1600 mg solution over 7 days under Fed conditions - Part C.

| Parameters | N | Cohort 1<br>1200 mg fed | N | Cohort 2<br>1600 mg fed |
| --- | --- | --- | --- | --- |
| C<sub>max</sub>, µg/mL | 4 | D1: 0.68 [1.57]<br>D7: 0.67 [1.26] | 4 | D1: 1.39 [1.61]<br>D7: 0.90 [1.22] |
| AUC<sub>last</sub>, µg*h/mL | 4 | D1: 2.48 [3.40]<br>D7: 3.97 [1.59] | 4 | D1: 8.18 [1.46]<br>D7: 4.34 [1.39] |
| t<sub>max</sub>, h | 4 | D1: 2.0 [1.0-4.0]<br>D7: 1.75[0.5-4.0] | 4 | D1: 1.75 [1.0-2.0]<br>D7: 1.5 [1.0-2.0] |
| C<sub>8h</sub>, µg/mL | 4 | D1: BQL D7: 0.14<br>[2.06] | 4 | D1: 0.41 [1.93]<br>D7: 0.16 [1.27] |
| C<sub>12h</sub>, µg/mL | 4 | D1: BQL D7: 0.10<br>[2.31] | 4 | D1: 0.16 [1.76]<br>D7: 0.08 [1.64] |
| C<sub>24h</sub>, µg/mL | 4 | D1: BQL D7: 0.05<br>[2.78] | 4 | D1: 0.03 [1.43]<br>D7: 0.03 [1.46] |
| t<sub>1/2</sub>, h | 3<br>2 | D1: 5.26 [1.06]<br>D7: 5.76 [1.01] | 4<br>3 | D1: 4.15 [1.11]<br>D7: 5.49 [1.24] |

C<sub>max</sub>: Max plasma concentration; AUC<sub>last</sub>: Area under plasma conc-time curve to last quantifiable conc; t<sub>max</sub>: Time to reach C<sub>max</sub> (median [range]); C<sub>8h</sub>, C<sub>12h</sub>, C<sub>24h</sub>: Plasma conc at 8, 12, 24 hrs post-dose; t<sub>1/2</sub>: Terminal half-life; n: Subjects; BQL: Below quantifiable limit. Geometric mean and standard deviation presented.

C_max_ (GM) between day 1 (0.68 µg/ml) to day 7 (0.67 µg/ml) in dose group 1200 mg was similar on both days. For dose group, 1600 mg C_max_ (GM) was higher overall than in the 1200 mg group but declined between day 1 (1.39 µg/ml) and day 7 (0.90 µg/ml).

AUC_last_ (GM) had increasing trends from day 1 (2.48 µg*h/ml) to day 7 (3.97 µg*h/ml) in dose group 1200 mg. In contrast, in the dose group 1600 mg, AUC_last_ (GM) decreased 47 % in the same interval yet exceeded the dose group 1200 mg 3.3-fold on day 1 (8.18 µg*h/ml) and was only slightly higher on day 7 (4.34 µg*h/ml).

Mean and individual trough concentrations of niclosamide measured on days 3 to 7 were all below the quantifiable limit for the 1200 mg and 1600 mg groups, suggesting no accumulation after multiple dosing.

The high variability, the poor bioavailability and the decrease in AUC after multiple dosing might all be connected to the side effects experienced from the PEG carrier solution. The correlation between adverse events and pharmacokinetics was therefore further analyzed.

#### Correlation between adverse events and plasma concentrations

From the AE summaries (Tables 4-6), after nausea, diarrhoea was the most frequent preferred term for AE classification. Diarrhoea impacts intestinal passage time and might therefore reduce absorption.

To analyze the relationship between exposure variability and diarrhoea further, we compared the subjects with diarrhoea and the subjects without diarrhoea to dose-normalized C_max_ and AUC, pooling the 1600 mg groups (A3 fed/fast, B, C) and 1200 mg Part C group. We considered AEs on day 1 for all groups, and additionally on day 7 for group C, because Cmax and AUC were only measured on these days. Groups A1 and A2 showed no diarrhoea on day 1. There is a pattern suggesting that subjects with diarrhoea had a lower C_max_ and AUC_last_ compared to subjects who did not experience diarrhoea after drug intake (Fig 3). However, even in subjects without diarrhoea, variability in exposure remained high with a roughly fourfold increase from lowest to highest value.

**Fig 3.**
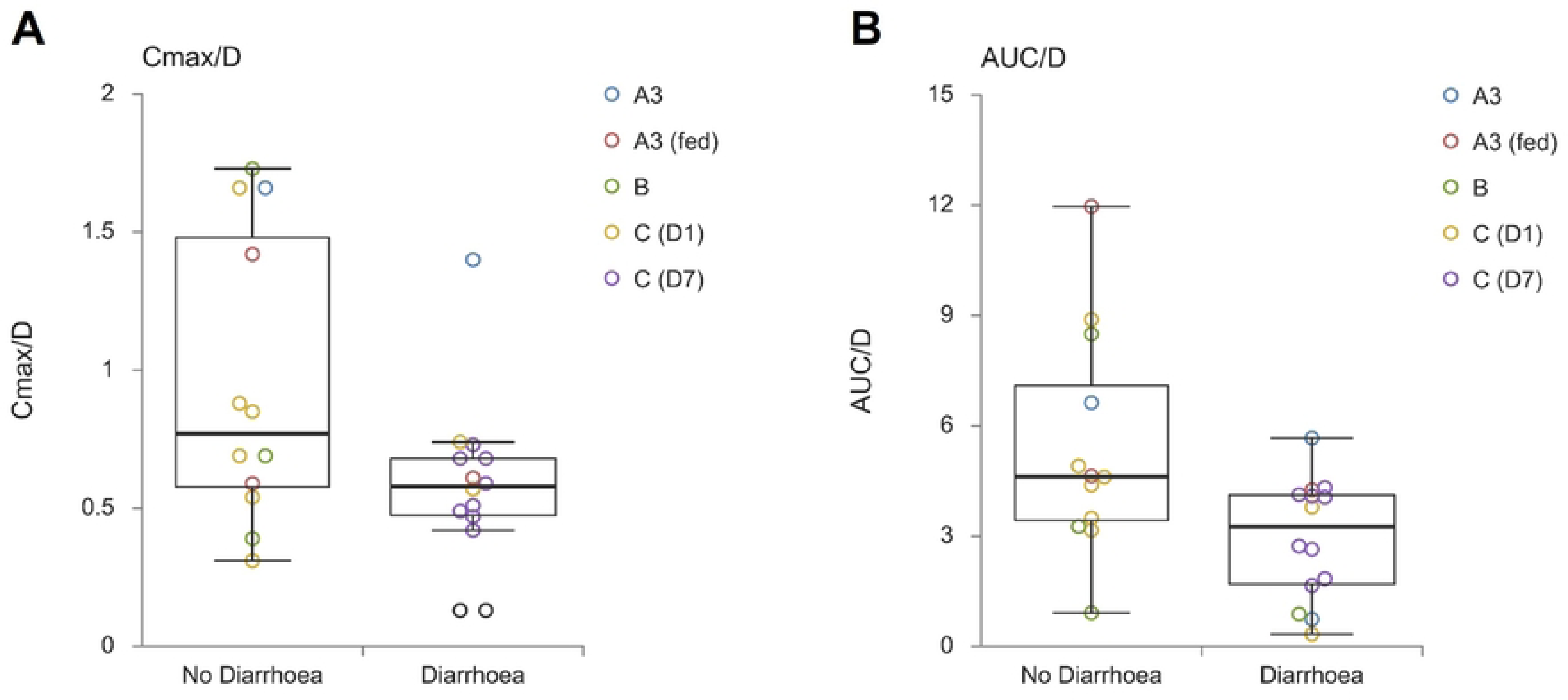
Diarrhoea Incidence vs. Dose-normalized Cmax (A) and AUC (B) in Niclosamide-treated Subjects. This figure presents the comparison of individual dose-normalized (A) Cmax (µg/mL per g) and (B) AUC (µg/mL per g) data in relation to the incidence of diarrhoea (no diarrhoea vs. diarrhoea) after niclosamide solution administration.

To inform the relationship between exposure and AEs, we plotted the number of any TEAE reported against a group of “high absorbers” and a group of “regular absorbers”. High absorption was here arbitrarily defined as being at least 2 µg/ml (Fig 4). Interestingly, we did not find a clear correlation between niclosamide exposure and TEAS. Two of the high absorbers did not report any adverse event and three high absorbers reported two adverse events which where all mild. Overall, the analysis suggests that number and severity of AEs is independent of the niclosamide exposure level reached in the individual.

**Fig 4.**
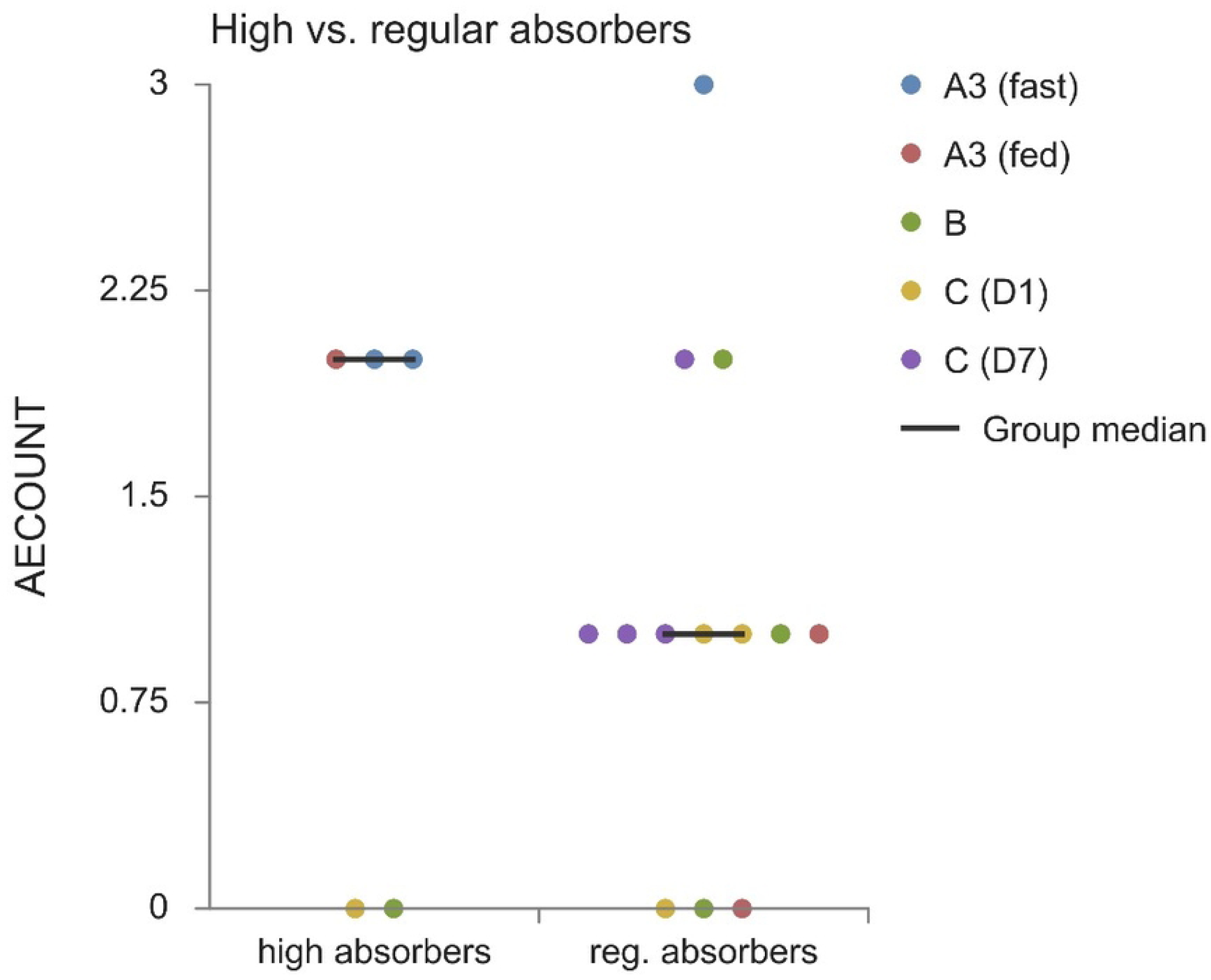
TEAEs Comparison between High Absorbers (Cmax > 2µg/ml) and Regular Absorbers (Cmax < 2µg/ml). This figure compares the number of treatment-emergent adverse events (TEAEs) in high absorbers and regular absorbers after niclosamide administration, highlighting the differences in TEAE distribution between the two groups.

## Discussion

The primary objective of this Phase 1 trial was to assess the safety and pharmacokinetics of a novel niclosamide solution. The study included a single ascending dose (SAD) consecutive group design (Part A) including assessment of the effect of food on oral absorption of niclosamide, a crossover comparison of bioavailability with a marketed chewing tablet (Part B), and a multiple ascending dose (MAD) design (Part C). Cohort A3 was repeated under fed conditions to study whether food was affecting systemic exposure of the niclosamide solution. The study protocol allowed for dosing in parts B and C to be performed under either fasting or fed conditions, depending on the bioavailability data obtained.

To evaluate the relative bioavailability of the niclosamide solution compared with the chewing tablet, a crossover part (Part B) was performed to exclude inter-individual differences in absorption and thus minimize the variability of PK parameters. The niclosamide solution’s highest tolerated and safe dose in part A was compared to the marketed chewing tablet at the highest approved dose of 2000 mg. Due to the higher exposure of niclosamide solution observed in cohort A3, the IMP was administered with prior food intake.

An MAD design (Part C) was chosen to investigate safety and PK after repeated drug administration. Since multiple gastrointestinal AEs were observed at the 1600 mg dose in the single dose parts, two different dose levels, 1600 mg, and 1200 mg, were chosen. Dosing in the 1600 mg could have been stopped if intolerable gastrointestinal AEs prevented further intake. In that case, dosing could have been continued with the 1200 mg niclosamide solution in these discontinued subjects, in addition to the four other subjects in the 1200 mg group.

In cohorts A and C, a placebo control was included that allowed differentiation between drug-related AEs and AEs that may occur without active treatment.

In 28 healthy volunteers, the investigational niclosamide solution was reasonably well-tolerated in doses up to 1600 mg, with no severe TEAEs reported. The overall frequency of suspected related TEAEs was 18 TEAEs in 7 (58.2%) subjects in part A, 3 TEAEs in 2 (50%) subjects in part B, and 28 TEAEs in 12 (100%) subjects in part C. The most common related TEAE were gastrointestinal symptoms such as diarrhoea (including liquid stool), nausea, and vomiting shortly after IMP administration. No trends were observed in hematologic and clinical chemistry parameters, vital signs, or ECG during the study.

TEAEs reported across the SAD and MAD parts were mild to moderate intensity. There were no discontinuations, and no TEAE needed medical intervention, except for “abdominal pain” in cohort A2, which was treated with oral ibuprofen. The proportion of gastrointestinal events increased with increasing dose and volume of solution, with no TEAEs in cohort A1, 66% in cohort A2, 100% in the fasted part of cohort A3, 67% in the fed part (2/3 subjects), 50% in part B after receiving solution and 100% in both treatment groups of part C. A relationship between TEAEs frequency and exposure was not found. Since subjects receiving placebo also had the same proportion of AEs and the frequency of AEs was independent of AUC or C_max_, but incidence increased with solution volume, the excipient PEG 400, an osmotic agent, likely led to these gastrointestinal events. This is corroborated by the fact that the volume of the excipient also increases with the dose of the drug. The volume of niclosamide solution was 5 ml for 200 mg, 15 ml for 600 mg, 30 ml for 1200 mg, 40 ml for 1600 mg, and 20 ml in part C for the placebo. PEG-400 was used here as a solvent to increase absorption in the intestine. However, PEG has an osmotic effect and is the basis of some laxatives used in gastroenterology, known to cause GI side effects that could affect absorption [15, 17].

Comparison of C_max_ and AUC_last_ versus dose revealed a non-linear increase in exposure over the dose range investigated, while deviation from dose proportionality was not evident between cohort A1 (200mg) and cohort A2 (600mg). This pattern could be explained by poor solubility, which could potentially limit the oral bioavailability of this niclosamide formulation at higher doses. At higher oral doses, the undissolved niclosamide might precipitate in some subjects. Furthermore, a positive food effect after a standard high-fat, high-calorie breakfast was observed; Fed GM C_max_ and AUC_last_ were 1.2-fold and 2-fold higher for fed compared to fasting conditions. Compared with fasting, the median t_max_ of mean plasma concentrations was prolonged under fed conditions, with individual plasma concentrations showing a delayed decline after C_max_ and measurable niclosamide plasma concentrations up to 12 hours after administration. It is noted, however, that the food effect might also – at least in part – be explained by the fewer observed gastro-intestinal AEs in the fed as compared to the fasted subjects.

Subsequent cohorts A3 and parts B and C were conducted under fed conditions, as the systemic exposure of the niclosamide solution was increased prior to food intake.

The variability of the PK parameters remained very high in all dose groups, especially under fasting conditions and at higher doses. The high variability could partly be explained by the gastrointestinal side effects like diarrhoea or vomiting which reduce resorption.

In part A, high variability in exposure was observed with even higher variability under fasting conditions and at increasing dose levels. The geometric coefficient of variation (Geo CV) for C_max_ in Cohort A1, A2, A3 fasted, and A3 fed, was 101%, 44%, 253%, and 53% and for AUC_last_ it was 8%, 58%, 185%, and 62%, respectively. After a high-fat,/high-calorie meal, PK variability was almost 2-fold lower for this treatment speaking for a dose recommendation under fed conditions in clinical practice. Variability was also high in part B, especially for the niclosamide solution. The Geo CV for C_max_ was 153% and 38%, and for the AUC_last_ was 152% and 20% for the solution and chewable tablets, respectively, making the chewing tablet more favourable for clinical use due to more predictable pharmacokinetics. In Part C, variability was less pronounced, decreasing from day 1 to day 7, with similar values in both dose groups, with Geo CV for C_max_ ranging from min. 20% to max. 60% and AUC_last_ ranging from min. 10% to max. 39%.

Contrary to our expectations, the novel formulation did not significantly improve intestinal absorption compared to the marketed chewing tablet. Data obtained from part B of our study indicate that the niclosamide solution may not hold a significant advantage over the chewing tablet with regard to bioavailability. However, it is important to consider that the FDA-standard breakfast consumed during the trial might have been excessively large, thereby also influencing absorption. This sizeable, high-fat meal can independently contribute to gastrointestinal side effects. Moreover, it is possible that bioavailability could be further optimized by adjusting the ratio of niclosamide to PEG, specifically by reducing the amount of PEG utilized, as it was likely causatively responsible for the observed gastro-intestinal side effects.

Following multiple, once-daily doses of 1600 mg over 7 days, AUC_last_ was 3,3-fold and 1,1-fold higher on day 1 and 7, respectively, compared to the 1200 mg group. Exposure decreased from day 1 to day 7 in the 1600 mg group, most strikingly with AUC_last_ decreasing by 47 %. Whereas 1200 mg AUC_last_ increased by 38%. Gastrointestinal symptoms occurred more frequently with multiple dosing than single dosing of parts A and B, possibly affecting niclosamide plasma levels. Following multiple doses QD over 7 days, niclosamide showed no accumulation.

There is a pattern suggesting that diarrhoea after study drug ingestion results in reduced absorption, as reflected by a lower C_max_ and AUC_last_ compared with subjects who did not have diarrhoea after drug administration (Fig 3). The mean concentration-time profiles of each cohort remained similar throughout the study, showing a rapid increase and decline in plasma concentration. There were individual high absorbers (with C_max_ values > 2 µg/ml) with the highest values for C_max_ and AUC_last_ of up to 2.77 µg/ml and 19.15 µg*h/ml, respectively, showing only mild TEAEs and no moderate or severe TEAEs. No clear pattern could be identified by comparing the counts of TEAEs from high absorbers with those from regular absorbers (Fig 4), indicating that high plasma levels of niclosamide do not result in acute toxicity which might encourage further exploration of pharmacologically active niclosamide exposure levels. Obviously, the small sample size of our study does not allow any firm conclusions on safe exposure levels.

Up to now, there is only scarce public information on the pharmacokinetics of niclosamide. A phase I study by M. T. Schweizer et al. of niclosamide as a chewing tablet in combination with enzalutamide in men with castration-resistant prostate cancer that enrolled 3 patients in a 500-mg TID cohort and 2 patients in a 1000-mg TID cohort for 4 weeks showed a niclosamide range for C_max_ from 0.04 to 0.18 µg/mL (Dose normalized: C_max/_D 0.07 to 0.36 µg/mL per g), for t_max_ from 1 to 6 h and for t_1/2_ from 1.27 to 5.61 h [18]. We showed a C_max/_D ranging from 0.13 µg/mL per g in the 1600 mg group to 4.90 µg/mL per g in the 200 mg group. Except for three individuals (RND: 132 in A3 fasted, 214 in B, and 319 in C on day 1), every C_max_ was higher in our study than in the aforementioned study. When only comparing the identical formulation (chewing tablet), it is noticeable that in our study Part B C_max/_D was still twice as high (C_max_/D 0.30 to 0.62 µg/mL per g) than in Schweizer et al.’s study. This could be due to the high-fat breakfast. Unfortunately, in the study published previously patients were allowed to take niclosamide fasting or non-fasting, so it cannot be clearly determined whether the high-fat breakfast resulted in the difference in PK.

Schweizer et al. showed that C_max_, AUC, and t_max_ exhibited large individual variability in both dose groups, which is confirmed by our study. Also, the C_max_ values determined in the study including cancer patients were lower than the range of 0.25-6.0 μg/ml described in the SmPC. Niclosamide acts on multiple highly conserved signalling pathways including, but not limited to, the inhibition of several inflammasomes [19], the inhibition of Signal transducer and activator of transcription protein 3 (STAT3) [20], the upregulation of autophagy via S-phase kinase-associated protein 2 (SKP2) inhibition [21], inhibition of TMEM16A [22] and the inhibition of the Wnt/β-catenin pathway [23]. Interfering with these signalling pathways could have many beneficial effects, but could also pose certain risks. Potential benefits and risks of interfering with these pathways are described in the public domain.

By inhibiting inflammasomes, the immune system may be compromised, which could lead to opportunistic infections caused by fungi and certain bacteria [24]. Similarly, inhibition of STAT3 could lead to humoral immunodeficiency, promoting the reactivation of viruses and impaired ability to fight infections [25].

Inhibition of SKP2 by niclosamide reduces the degradation of Beclin1, which promotes autophagic flux. Although there is evidence that activation of autophagy may limit tumor development, there is also suggestive evidence that inhibition of autophagy restricts the growth of preexisting tumours and enhances the response to tumor therapies [26]. Furthermore, Beta cells in human type 2 diabetes show signs of increased autophagic flux, which may promote the loss of beta cell mass [27]. Increased autophagy could also be harmful in certain cardiovascular diseases and contribute to disease progression suggesting that modulations of autophagic flux need to be well balanced [28].

Numerous therapeutic strategies are being developed to inhibit the Wnt/β-catenin pathway, which may be effective against several malignancies [29]. Activation and overexpression of this pathway are frequently observed in malignant tumours. However, the Wnt/β-catenin pathway also regulates embryonic development and tissue homeostasis in adulthood, maintaining bone, hair, skin, gastrointestinal tract, liver, and lung, and is involved in hematopoiesis and neurogenesis in adulthood [30, 31]. Expected adverse effects of inhibition of this pathway could include, for example, wound healing disorders, bone loss or fractures, hair loss, elevated liver enzymes, and anaemia.

However, none of these potential side effects were observed in our study, but they should be considered in future studies that might involve prolonged use of niclosamide.

A limitation of our study is the low number of subjects, which restricts the generalizability of the PK parameters. Our study was conducted under the circumstances of the COVID-19 pandemic, which necessitated expedited timelines to explore potential antiviral treatments and explains the lower number of subjects. Moreover, interindividual variability was high, limiting the interpretation of results. Furthermore, a notable limitation is the lack of pharmacodynamic and surrogate parameters, as the study was conducted in healthy volunteers rather than a target patient population. Consequently, assessing the potential therapeutic effect and underlying mechanisms of action of the novel formulation in treating malignancies or viral infections remains challenging.

However, even with a higher number of subjects, the argument would not change. Based on the existing number of subjects, the study offers a reasonable evaluation of the niclosamide solution. The results suggest that it has not demonstrated clear benefits over the tablet form in aspects such as bioavailability, safety, tolerability, and controllability.

Overall, this study showed that administering the niclosamide solution up to 1600 mg for 7 days QD was reasonably well-tolerated and exhibited an acceptable safety profile in healthy subjects. While diarrhoea and vomiting were not considered clinically relevant in healthy subjects and were not otherwise of concern to the DSMB, it could likely pose patients with serious conditions at risk because of exsiccosis, electrolyte disturbances, or impaired absorption of essential concomitant medications; thus, the risk would outweigh the benefit. Moreover, contrary to our expectations, the new formulation did not improve niclosamide absorption. The variability in exposure remained high, and sufficiently higher plasma concentrations were not achieved. In fact, according to our data, the chewable tablet is probably equally efficient to reach systemic exposure with fewer adverse effects with similar exposure.

## Conclusions

This study added to the understanding of niclosamide pharmacokinetics in humans. The tested niclosamide solution was reasonably well-tolerated, but bioavailability did not meet expectations, indicating that the niclosamide solution is not superior to treat systemic conditions compared to the established one. Consequently, the data do not support further clinical investigation of this particular niclosamide solution, but two key findings resulted and could have important implications for further clinical research with niclosamide.

First, individual high absorbers tolerated the study drug well, suggesting that potentially pharmacologically active plasma levels of niclosamide do not lead to acute toxicity, which might be relevant for potential dose escalation in subsequent studies. Secondly, a positive food effect of up to 2-fold after a standardized high-fat breakfast was observed.

Potential applications for niclosamide through multiple drug mechanisms addressing multiple pathways and drug targets show the broad spectrum of potential applications for this drug and underline the need for novel formulations of niclosamide.

## Data Availability

All relevant data are within the manuscript and its Supporting Information files.

## Supporting information

**S1 Table.** Summary of individual niclosamide PK parameters and Adverse Events.

**S1 CONSORT** checklist.

**S1 Protocol.** Study protocol.

## Acknowledgments

The authors would like to thank the volunteers participating in the study as well as their clinical and nursing staff at the Charité Research Organisation GmbH.

